# Headache Assessment Via Digital Platform in United States (Head-US): Baseline Study Characteristics, Diagnosis, and Treatment Patterns

**DOI:** 10.1101/2025.04.03.25325136

**Authors:** Ali Ezzati, Kristina M. Fanning, Devin Teichrow, Alexandre Urani, François Cadiou, Richard B. Lipton

## Abstract

**Objectives:** To summarize the baseline methods for the Headache Assessment Via Digital Platform in United States (Head-US) study and report accuracy of self- reported diagnosis of migraine on the platform, and patterns of acute and preventive treatment use among participants with migraine.

**Background:** The HeAD-US is a new, ongoing cohort study of patients with migraine in the US, centered around comprehensive data collection via the Migraine Buddy platform, a smartphone app. Data from such platforms can be used to provide real-world insights, capturing patients’ voices directly.

**Methods:** Participants from the Migraine Buddy app completed baseline surveys assessing demographics, headache characteristics, and treatment use. Migraine diagnosis was determined using the AMS/AMPP diagnostic module, and treatment effectiveness was evaluated using the migraine Treatment Optimization Questionnaire (mTOQ-6, defined as scores ≥ 6 indicating moderate-to-maximum efficacy).

**Results:** Of 6810 participants who completed the baseline questionnaire, 6267 (92.0%) met migraine criteria and were included in the rest of the study. Participants had an average age of 41.5 years (SD=13.1, range 18-88), 5692 (90.8%) were female, 3924 (62.6%) met criteria for episodic migraine and 2343 (37.4%) met criteria for chronic migraine. Of the participants, 52.6% were using acute over-the-counter (OTC) medications, 73.2% were using acute prescription medications, 2.4% were using medical devices, and 60.1% were on preventive treatments. There was no significant difference in treatment effectiveness between participants on polytherapy (37.6%) and those on monotherapy (39.5%, p = 0.168). Among the 2621 patients on monotherapy with acute medications, the most common categories were triptans (39.0%), OTCs (28.2%), and gepants (20.8%). For patients on prescription monotherapy, individuals using gepants reported the highest rate of effective treatment (53.3%) in head-to-head comparison with triptans (47.28%, p=0.036), and opioid/barbiturates (27.1%, p<0.001).

**Conclusions:** HeAD-US participants have higher rates of chronic migraine and headache-related disability in comparison with large-scale epidemiologic studies. The HeAD-US study offers a valuable opportunity to leverage real-world data for understanding migraine patients, their treatment patterns, and outcomes.

## INTRODUCTION

Migraine is a prevalent and disabling neurological disorder affecting over one billion individuals worldwide. Given its high prevalence and significant individual and societal burden, migraine is ranked by the World Health Organization as the second most disabling neurological condition after stroke, contributing 45.1 million disability-adjusted life years globally.^1^ Accurate diagnosis and effective management of migraine remain among critical public health challenges.^2^ While the epidemiology of migraine has been extensively studied in the US and globally, understanding the evolving dynamics of consultation, diagnosis, and treatment patterns remains crucial. Over the past two decades, the way patients consume health-related information and provide data to researchers or clinicians has transformed dramatically, with digital platforms becoming the dominant medium. These digital health platforms have emerged as promising tools for real-world data collection, enabling large-scale studies on migraine characteristics, diagnostic accuracy, and treatment patterns. In the long run, these platforms have the potential to inform interventions targeting patient behavior and treatment strategies, ultimately improving health outcomes.^3^

Findings from prior national surveys, including the American Migraine Prevalence and Prevention (AMPP) Study, the Chronic Migraine Epidemiology and Outcomes (CaMEO) study, the Migraine in America Symptoms and Treatment (MAST) Study, and the OVERCOME study underscore the public health burden of migraine.^4-7^ The AMPP Study highlighted the underdiagnosis and undertreatment of migraine attacks, noting that only 56.2% of individuals with migraine had ever received a medical diagnosis, with many never receiving medical care from a clinician for their headaches and solely relying solely on over-the-counter medications.^6^ Similarly, the MAST Study revealed that while most individuals with migraine use acute treatments, preventive treatment remains underutilized, especially among men and those with fewer healthcare consultations.^4^

The Headache Assessment via Digital Platform in the United States (HeAD-US) study was designed to assess current patterns of symptomatology, consultation, diagnosis, and treatment in a large sample of people with migraine in the US who use digital platforms for recording their migraine symptoms and management. The HeAD-US leverages the Migraine Buddy application, a widely used smartphone-based platform for headache tracking and management. Previous research has established the feasibility of using similar digital platforms for migraine-related studies.^8, 9^ However, the HeAD-US study is unique in its scale, comprehensiveness, and focus on U.S. participants.

This manuscript presents baseline, first-year findings from the HeAD-US study, focusing on diagnostic accuracy and patterns of acute and preventive treatment among individuals meeting International Classification of Headache Disorders, Third Edition (ICHD-3) criteria for migraine. In the past, Migraine Buddy and similar platforms have been criticized for not verifying migraine diagnosis among participants. In the current report, we evaluate migraine diagnosis among app users using a validated diagnostic survey. In addition, we evaluate the real-world patterns and effectiveness of commonly used treatments and assessing its relations with patient-reported outcomes, this study aims to fill critical knowledge gaps in migraine management. The insights gained can inform clinical practice and guide future research on optimizing migraine care.

## METHODS

### Study design and participants

The HeAD-US is a cohort study of adults in the United States with migraine, surveyed through the Migraine Buddy application. Migraine Buddy is a smartphone app available on all platforms via Google Play and the Apple Store. Individuals with headaches self-download this app and use it for various purposes, including keeping a headache diary and obtaining summary reports about their headache patterns. Migraine Buddy has been used for conducting research studies in other formats in the past,^10-12^ however this is the first usage of this application to establish and follow a panel of people with migraine. Starting in September 2023, over approximately three months, users of the Migraine Buddy app were notified about the HeAD-US study and invited to participate. Participation was voluntary, and respondents did not receive an honorarium or any other form of compensation. All participants provided electronic consent for participation in a general, survey-based migraine study and subsequently received the study questionnaire through the Migraine Buddy application. All study procedures were approved by the Institutional Board Review.

The questionnaire included standard questions on sociodemographic information, headache features such as severity and frequency, neuropsychiatric symptoms, use of acute and preventive treatments, and treatment responses. Quality control process included a quality control question, (also known as attention checks), which was used in the middle of a survey, and through verifying programmed response ranges and performing consistency checks. Response options like “prefer not to answer,” “don’t know,” “does not apply to me,” and “don’t remember” were included to accommodate participants who couldn’t or didn’t want to provide a definitive answer to specific questions, aiming to minimize missing data. Survey questions were presented in the same order to all participants, prioritizing essential questions. Adaptive question logic was applied where appropriate.

A total of 6810 participants who completed the questionnaire were eligible for this study.

### Study measures

Study measures can be broadly categorized into the following groups:

1. Socioeconomic measures: Age, gender, marital status, and health insurance status.
2. Headache features and symptoms: Includes headache pain intensity, Migraine Symptom Severity Score (MSSS), ^13^ average monthly headache days, and cutaneous allodynia. We included the validated AMS/AMPP Diagnostic Module to assess the proportion of participants who used the app who met criteria for migraine.^13^
3. Depression and disability: Assessed using Depression and Anxiety (PHQ-4)^14^ and Perceived Stress Scale (PSS-4).^15^
4. Treatment patterns: Respondents reported their lifetime and current use of acute and preventive medications for migraine available at the time of the survey. Participants were asked separately about the classes of medications they use for acute or chronic treatment of migraine. Medication categories were provided, each listing several names of most common medications in each category, to capture the pharmacologic classes comprehensively.
5. Treatment outcomes: The Migraine Treatment Optimization Questionnaire-6 (mTOQ-6) evaluates the effectiveness and optimization of migraine treatments through six questions. This tool helps healthcare providers assess patient responses to medications and identify areas for improvement. Like other recall-based questionnaires, mTOQ-6 is subject to subjective self-report and recall bias. However, its items demonstrate high test-retest reliability and validity in clinical settings.^16^

### Establishing Migraine Diagnosis

All participants were asked to complete the AMS/AMPP diagnostic module.^13^ Episodic migraine (EM; <15 headache days/month) and chronic migraine (CM; ≥15 headache days/month) cases were identified based on the ICHD criteria.^17^

### Definition of Treatment Effectiveness

Treatment effectiveness was assessed using the Migraine Treatment Optimization Questionnaire (mTOQ-6), a validated tool for evaluating the efficacy of acute migraine treatments. The mTOQ-6 scores range from 0 to 8, with higher scores indicating greater perceived treatment effectiveness. For the purposes of this study, treatment effectiveness was defined as an mTOQ-6 score of 6 or higher, corresponding to “Moderate” (scores 6–7) or “Maximum” (score 8) treatment efficacy. Participants with scores below 6 were categorized as experiencing “Very Poor” (score 0) or “Poor” (scores 1–5) treatment efficacy.

### Statistical Analysis

Descriptive statistics were calculated for all study measures and reported as means with standard deviations for continuous variables and frequencies with percentages for categorical variables. Chi-square tests were used to examine group differences for categorical variables, while independent *t*-tests were employed for continuous variables. Statistical significance was set at *p* < 0.05, with test statistics and corresponding *p*-values reported for all comparisons.

To identify predictors of acute medication use, logistic regression analyses were conducted. The dependent variable in each model was the type of acute medication, and the independent variables included MIDAS score (categorized as none-to-mild vs. moderate-to-severe), age (continuous), sex (binary), and race (categorical). Post-hoc pairwise comparisons were performed to further explore differences across monotherapy classes with respect to MIDAS scores, PHQ-4 anxiety and depression scores, and MSSS scores.

Statistical analysis was performed with *SPSS Statistics for Macintosh, version 29*.*0; IBM, Armonk, NY*.

## RESULTS

### Migraine Diagnosis

We evaluated validity of diagnosis of migraine in participants based on the AMS/AMPP diagnostic module.^13^ Of all survey responders, 6267 participants (92.0%) met the ICHD-3 criteria for migraine, while an additional 339 participants (5.0%) met criteria for probable migraine. Furthermore, 97.0% (6606/6810) of users met an ICHD based case definition for probable or definite migraine. Among those meeting ICHD-3 migraine diagnostic criteria, (5932/6267) 94.7% had Self-reported medical diagnosis, compared to (483/543) 89.0% among those who did not meet the criteria. The rest of study analysis was restricted to individuals who met ICHD-3 migraine diagnostic criteria based on the AMS/AMPP diagnostic module.

#### Sample Characteristics

A total of 6267 participants currently met the ICHD-3 criteria for migraine and were eligible for this study. Participants had an average age of 41.5 years (SD=13.1, range 18-88), 90.8% were female, 3924 (62.6%) met criteria for EM and 2343 (37.4%) met criteria for CM. Table 1 summarizes the study sample characteristics for the whole sample with ICHD-3 diagnosis of migraine and the sample stratified by EM or CM diagnosis.

**Table 1.** Sample characteristics for the entire sample and by episodic and chronic migraine status: The HeAD Study, Sep. to Nov. 2023.

|  | **Episodic Migraine** | **Chronic Migraine** | **Total** |
| --- | --- | --- | --- |
| **N (%)** | 3924 (62.6%) | 2343 (37.4%) | 6267 (100.0%) |
| **Age, Mean (SD)** | 41.1 (12.8) | 42.3 (13.5) | 41.5 (13.1) |
| **Sex, Female** | 3553 (90.5%) | 2139 (91.3%) | 5692 (90.8%) |
| **Race, White or Caucasian** | 3318 (84.6%) | 1981 (84.5%) | 5299 (84.6%) |
| **Race, Black or African American** | 117 (3.0%) | 67 (2.9%) | 184 (2.9%) |
| **Ethnicity, Hispanic** | 264 (6.7%) | 140 (6.0%) | 404 (6.4%) |
| **Race, White or Caucasian (Non-Hispanic)** | 3169 (80.8%) | 1900 (81.1%) | 5069 (80.9%) |
| **Race, Black or African American (Non-Hispanic)** | 109 (2.8%) | 61 (2.6%) | 170 (2.7%) |
| **Using Preventive therapy** | 2056 (52.4%) | 1711 (73.0%) | 3767 (60.1%) |
| **Not using acute or preventive medications** | 404 (10.3%) | 171 (7.3%) | 575 (9.2%) |
| **Not using any acute medication** | 574 (14.6%) | 346 (14.8%) | 920 (14.7%) |
| **Migraine Symptom Severity Score** | 17.9 (2.2) | 18.3 (2.1) | 18.1 (2.2) |
| **Disability (MIDAS)** |  |  |  |
| None to Mild *grade I (0-5) to grade II (6-10)* | 479 (12.2%) | 55 (2.3%) | 534 (8.5%) |
| Moderate *grade III (11-20)* | 663 (16.9%) | 67 (2.9%) | 730 (11.6%) |
| Severe *grade IV (≥21)* | 2782 (70.9%) | 2221 (94.8%) | 5003 (79.8%) |
| **Anxiety Symptoms (PHQ4)** |  |  |  |
| Absent | 2404 (61.3%) | 1197 (51.1%) | 3601 (57.5%) |
| Present | 1519 (38.7%) | 1146 (48.9%) | 2665 (42.5%) |
| **Depressive Symptoms (PHQ4)** |  |  |  |
| Absent | 2946 (75.1%) | 1447 (61.8%) | 4393 (70.1%) |
| Present | 978 (24.9%) | 896 (38.2%) | 1874 (29.9%) |

### Patterns of Current Preventive Medication Use

Table 2 presents patterns of preventive medication use stratified by episodic migraine (EM) and chronic migraine (CM) status. Across the entire sample, 25.1% of participants reported using anti-CGRP monoclonal antibodies, with significantly higher use in those with CM (32.7%) compared to those with EM (20.5%, p<0.001). Similarly, antidepressant use was greater among CM participants (27.0%) compared to EM (16.8%, p<0.001), while anti-seizure medications were used by 24.2% of CM participants versus 15.5% of EM participants (p<0.001). Beta blockers, Botox, and gepants followed the same trend, with higher usage among CM participants (20.9%, 27.1%, and 19.5%, respectively) compared to EM participants (13.8%, 10.1%, and 10.4%, all p<0.001).

**Table 2.** Patterns of current preventive medication use for the entire sample and by episodic and chronic migraine status, Sept to Nov, 2023.

|  | **Episodic Migraine** | **Chronic Migraine** | **Total** | **Chi<sup>2</sup>** | **p-value** |
| --- | --- | --- | --- | --- | --- |
| **N (%)** | 3924 (62.6%) | 2343 (37.4%) | 6267 (100.0%) |  |  |
| **Anti-CGRP monoclonal antibodies** | 804 (20.5%) | 767 (32.7%) | 1571 (25.1%) | 117.134 | <0.001 |
| **Anti-depressants** | 660 (16.8%) | 632 (27.0%) | 1292 (20.6%) | 92.430 | <0.001 |
| **Anti-seizure medications** | 608 (15.5%) | 566 (24.2%) | 1174 (18.7%) | 72.314 | <0.001 |
| **Beta blockers** | 540 (13.8%) | 489 (20.9%) | 1029 (16.4%) | 54.028 | <0.001 |
| **Botox** | 396 (10.1%) | 636 (27.1%) | 1032 (16.5%) | 310.144 | <0.001 |
| **Gepants** | 410 (10.4%) | 456 (19.5%) | 866 (13.8%) | 100.087 | <0.001 |

Table 3 summarizes patient-reported outcomes based on preventive treatment use. Among all participants, 60.1% were currently using preventive treatment, while (4102/6267) 65.5% reported *ever* having used migraine preventive treatment. Current preventive medication use was less likely among individuals with no-to-mild disability (p<0.001), no depressive symptoms (p=0.019), episodic migraine (p<0.001), or lower MSSS scores (p<0.001).

**Table 3.** Patient-reported outcomes by Preventive Treatment Use.

|  | No Current Preventive use | Current Preventive Use | Total | Chi<sup>2</sup> | p-value |
| --- | --- | --- | --- | --- | --- |
| **N (%)** | 2500 (39.9%) | 3767 (60.1%) | 6267 (100.0%) |  |  |
| **Disability (MIDAS)** |  |  |  |  |  |
| None to Mild | 308 (12.3%) | 226 (6.0%) | 534 (8.5%) | 138.468 | <0.001 |
| Moderate | 376 (15.0%) | 354 (9.4%) | 730 (11.6%) |  |  |
| Severe | 1816 (72.6%) | 3187 (84.6%) | 5003 (79.8%) |  |  |
| **Anxiety Symptoms (PHQ4)** |  |  |  |  |  |
| Absent | 1435 (57.4%) | 2166 (57.5%) | 3601 (57.5%) | 0.004 | 0.952 |
| Present | 1064 (42.6%) | 1601 (42.5%) | 2665 (42.5%) |  |  |
| **Depressive Symptoms (PHQ4)** |  |  |  |  |  |
| Absent | 1794 (71.8%) | 2599 (69.0%) | 4393 (70.1%) | 5.485 | 0.019 |
| Present | 706 (28.2%) | 1168 (31.0%) | 1874 (29.9%) |  |  |
| **EM vs CM** |  |  |  |  |  |
| EM | 1868 (74.7%) | 2056 (54.6%) | 3924 (62.6%) | 260.403 | <0.001 |
| CM | 632 (25.3%) | 1711 (45.4%) | 2343 (37.4%) |  |  |
|  | **Mean (SD)** | **Mean (SD)** | **Mean (SD)** | **F** | **p-value** |
| **Migraine Symptom Severity Score** | 17.9 (2.3) | 18.2 (2.2) | 18.1 (2.2) | 20.017 | <0.001 |
| **Note:** Disability measured by MIDAS score, Anxiety and depressive symptoms are measured by PHQ4 scores. Abbreviations: EM=Episodic Migraine; CM=Chronic Migraine; MSSS= Migraine Symptom Severity Score |  |  |  |  |  |

### Patterns of Current Acute Treatment Use

Only (920/6267) 14.7% of surveyed migraine patients were not currently treating their migraine attacks with any acute treatment. Among the total sample, 52.6% were using OTC medications, (4620/6267) 73.2% were using prescription medications (2572/6267) 84.9% were using both OTC and prescription medications, and (152/6267) 2.4% were using medical devices. Of the medical device users, (147/152) 96.7% also used OTC or prescription medications.

Chronic migraine (CM) respondents were more likely than episodic migraine (EM) respondents to use OTC medications (54.5% vs. 51.5%, p<0.05), Opioids or Barbiturates (14.5% vs. 7.9%, p<0.001) and Gepants (35.0% vs. 25.9%, p<0.001). However, CM respondents were less likely to use triptans compared to EM respondents (47.5% vs. 51.6%, p<0.01). Additionally, participants with CM were significantly more likely to use medical devices (4.0% vs. 1.5%, p<0.001). These patterns are detailed in Table 4.

**Table 4.** Patterns of Acute medication use for the entire sample and by episodic and chronic migraine status.

|  | **Episodic**<br>**Migraine** | **Chronic**<br>**Migraine** | **Total** | **Chi<sup>2</sup>** | **p-value** |
| --- | --- | --- | --- | --- | --- |
| **N (%)** | 3924 (62.6%) | 2343 (37.4%) | 6267 (100.0%) |  |  |
| **Acute Medication Use – Monotherapy or Polytherapy\*** |  |  |  |  |  |
| OTCs or NSAIDs | 2022 (51.5%) | 1277 (54.5%) | 3299 (52.6%) | 5.204 | 0.023 |
| Opioids or Barbiturates | 311 (7.9%) | 339 (14.5%) | 650 (10.4%) | 67.562 | <0.001 |
| Triptans | 2024 (51.6%) | 1112 (47.5%) | 3136 (50.0%) | 9.958 | 0.002 |
| Gepants | 1017 (25.9%) | 819 (35.0%) | 1836 (29.3%) | 57.851 | <0.001 |
| **Acute Medication Use - Monotherapy** | 1648 (42.0%) | 773 (33.0%) | 2421 (38.6%) | 50.191 | <0.001 |
| OTCs or NSAIDs | 516 (13.1%) | 222 (9.5%) | 738 (11.8%) | 19.069 | <0.001 |
| Opioids or Barbiturates | 40 (1.0%) | 45 (1.9%) | 85 (1.4%) | 8.906 | 0.003 |
| Triptans | 738 (18.8%) | 284 (12.1%) | 1022 (16.3%) | 48.052 | <0.001 |
| Gepants | 354 (9.0%) | 222 (9.5%) | 576 (9.2%) | .362 | 0.548 |
| **Medical Device Use** |  |  |  |  |  |
| Any Medical Device | 59 (1.5%) | 93 (4.0%) | 152 (2.4%) | 37.688 | <0.001 |
| Cefaly Supraorbital TENS | 39 (1.0%) | 61 (2.6%) | 100 (1.6%) | 24.206 | <0.001 |
| electroCore nVS External vagal nerve stimulation | 2 (0.1%) | 4 (0.2%) | 6 (0.1%) | 2.200 | 0.138 |
| eNeura TMS single pulse transcutaneous magnetic stimulation | 6 (0.2%) | 8 (0.3%) | 14 (0.2%) | 2.340 | 0.126 |
| Nerivio Armband Remote electrical neuromodulation | 20 (0.5%) | 29 (1.2%) | 49 (0.8%) | 10.024 | 0.002 |

**Note.** \* Total number of participants who reported use of one or multiple treatments for migraine. OTC = over the counter; NSAIDs = Nonsteroidal anti-inflammatory drugs; TMS = transcranial magnetic stimulation; TENS = transcutaneous electrical nerve stimulator; nVS = noninvasive vagus nerve stimulator

Among those on acute monotherapy for treatment of acute attacks, the majority (1022/2421, 42.2%) treated their attacks with triptans, followed by OTCs (738/2421, 30.5%) and Gepants (576/2421, 23.8%). We compared the impact of headache characteristics, migraine disability, and comorbidities on acute medication use patterns (**Table 5**).

**Table 5.** Headache Characteristics, Comorbidities, & Outcomes in Patients on Monotherapy for Migraine by Acute Medication Class.

|  | NSAID or OTC | Opioid or Barbiturate | Triptans | Gepants | Total | P value* |
| --- | --- | --- | --- | --- | --- | --- |
| **N (%)** | 738 (11.8%) | 85 (1.4%) | 1022 (16.3%) | 576 (9.2%) | 2421 (38.6%) |  |
| **Age, Mean (SD)** | 36.5 (12.6) | 50.2 (13.7) | 44.0 (13.4) | 42.4 (12.3) | 41.5 (13.4) | <0.001 |
| **Female Sex** | 667 (90.4%) | 78 (91.8%) | 906 (88.6%) | 535 (92.9%) | 2186 (90.3%) | 0.124 |
| **Race, White or Caucasian** | 586 (79.4%) | 74 (87.1%) | 891 (87.2%) | 480 (83.3%) | 2031 (83.9%) | <0.001 |
| **Race, Black or African American** | 29 (3.9%) | 5 (5.9%) | 13 (1.3%) | 26 (4.5%) | 73 (3.0%) |  |
| **Ethnicity, Hispanic** | 66 (8.9%) | 6 (7.1%) | 58 (5.7%) | 38 (6.6%) | 168 (6.9%) | 0.234 |
| **Monthly Headache Days** | 10.3 (8.0) | 14.2 (8.6) | 10.2 (7.5) | 12.1 (8.4) | 10.8 (7.9) | <0.001 |
| **Migraine Symptom Severity Score Mean (SD)** | 17.9 (2.3) | 18.1 (2.0) | 18.0 (2.3) | 18.0 (2.2) | 18.0 (2.3) | 0.405 |
| **Disability (MIDAS)** |  |  |  |  |  |  |
| None to Mild | 65 (8.8%) | 5 (5.9%) | 109 (10.7%) | 49 (8.5%) | 228 (9.4%) | 0.124 |
| Moderate | 106 (14.4%) | 7 (8.2%) | 137 (13.4%) | 63 (10.9%) | 313 (12.9%) |  |
| Severe | 567 (76.8%) | 73 (85.9%) | 776 (75.9%) | 464 (80.6%) | 1880 (77.7%) |  |
| **Anxiety symptoms (PHQ4)** |  |  |  |  |  |  |
| Absent | 380 (51.5%) | 54 (63.5%) | 645 (63.1%) | 329 (57.1%) | 1408 (58.2%) | <0.001 |
| Present | 358 (48.5%) | 31 (36.5%) | 377 (36.9%) | 247 (42.9%) | 1013 (41.8%) |  |
| **Depressive symptoms (PHQ4)** |  |  |  |  |  |  |
| Absent | 497 (67.3%) | 56 (65.9%) | 758 (74.2%) | 407 (70.7%) | 1718 (71.0%) | 0.012 |
| Present | 241 (32.7%) | 29 (34.1%) | 264 (25.8%) | 169 (29.3%) | 703 (29.0%) |  |
| **Note:** MIDAS = Migraine Disability Assessment; mTOQ = Migraine Treatment Optimization Questionnaire; ICHD-3 = International Classification of Headache Disorders, Third Edition. |  |  |  |  |  |  |

Patients with moderate to severe disability (MIDAS) were more likely to use acute treatments including OTCs (OR = 1.583, CI = 1.320-1.898, *p* < 0.001), Opioids/Barbiturates (OR = 2.506, CI = 1.656-3.791, *p* < 0.001), and Gepants (OR = 1.330, CI = 1.063-1.665, *p* = 0.013) than patients with mild disability. However, triptan use was not significantly related to MIDAS disability (OR = 1.102, CI = 0.920-1.322, *p* = 0.292). Interpretation of these findings is provided in the discussion.

OTC use was similar between patients with (53.8%) and without (52.1%) depressive symptoms (p=0.214). Opioid/Barbiturate use was higher among patients with depressive symptoms (11.4%) compared to those without (9.9%) (p=0.076). Gepant use was lower among patients with depressive symptoms (27.7%) compared to those without (30.0%) (p=0.079). Triptan use was significantly lower among patients with depressive symptoms (46.7%) compared to those without (51.5%) (p<0.001).

The presence of anxiety symptoms was significantly associated with higher rates of OTC use (54.8% vs 51.0%, p=0.003) and lower rates of Triptan use (47.6% vs 51.9%, p<0.001). Opioid/Barbiturate use rates were similar between anxiety symptoms present and absent (10.4% vs 10.4%, p=0.970). Gepant use was lower in the presence of anxiety symptoms than the absence of anxiety symptoms (28.1% vs 30.2%, p=0.083).

### Treatment Efficacy of Acute Treatments for Migraine based on mTOQ Scores

Treatment effectiveness, as defined by mTOQ-6 scores ≥ 6, did not significantly differ between participants using polytherapy (37.6%) and those on monotherapy (39.5%, p = 0.168). Among participants using monotherapy, gepants demonstrated the highest effectiveness (53.3%) compared to triptans (47.8%, p = 0.036) and opioid/barbiturate users (27.1%, p < 0.001). After classifying participants into EM and CM categories, gepants consistently demonstrated the highest rate of effective treatment in each category.

## DISCUSSION

Migraine Buddy and similar apps provide a powerful and efficient method for collecting data from a large population of individuals living with migraine. Historically, app-based studies have faced criticism regarding diagnostic uncertainty, raising questions about the reliability of self-reported migraine diagnoses. However, the baseline results from the first wave of the HeAD-US study address this concern, showcasing essential aspects of migraine diagnosis and management within a large U.S.-based cohort using data from a digital platform. Notably, 97% of participants met ICHD-3 criteria for migraine or probable migraine, and 92.0% met criteria for migraine, demonstrating high diagnostic accuracy. To our knowledge, this is the first study to validate the reliability of large-scale smartphone-based headache platforms for real-world migraine diagnosis. These tools not only enable robust data collection but also provide opportunities for feedback and enhancing patient care. Additionally, the approach used here illustrates an effective method for identifying individuals who meet migraine criteria using the AMS/AMPP diagnostic module, further solidifying the potential of digital health solutions in migraine research and management.

The patterns of preventive medication use differ substantially among the HeAD-US, OVERCOME, AMPP, and MAST studies, reflecting variations in recruitment methods, participant demographics, and treatment access. Other studies have shown that less than half of participants using prescription medications for acute migraine treatment.^6, 7^ They have also highlighted the underuse of preventive treatments among migraine patients.^4^ In the HeAD-US cohort, 60.1% of participants were currently using preventive treatments, with anti-CGRP monoclonal antibodies and Botox being the most common classes. This difference may reflect the HeAD-US study’s digital recruitment methodology, which likely attracted individuals with more severe or treatment-resistant migraine. A strength of the app-based study is that we identify people much more likely to be seeking treatment. The disadvantage is that we are less likely to enroll people with less severe migraine for whom better migraine outcomes may not be a priority. The broader, demographically representative samples in population studies provide a sharper focus on the entire spectrum of migraine. We hypothesize that the users of apps, may represent a motivated group, seeking better outcomes amenable to studies and perhaps changes in their approaches to treatment. Even in the HeAD study there is an apparent continued underuse of both acute and preventive treatments raises concerns about potential barriers to care, including medication access, cost, and treatment adherence.

Furthermore, substantial differences exist in patterns of acute medication use among the HeAD-US, OVERCOME, AMPP, MAST, and CaMEO studies, which underscore the evolving treatment landscape and access disparities over the past two decades. The OVERCOME study reported relatively lower triptan use, with less than one-quarter of participants using this class of medications and very few adopting newer therapies like gepants. The latter is no surprise given that gepants we considered a very new class of medications at the time of conduct of OVERCOME study. A significant majority relied primarily on OTC options. Similarly, the AMPP study found that nearly half of participants exclusively used OTC medications for acute symptom management, while only about one-fifth used prescription treatments. The MAST study highlighted a similar reliance on OTC medications, with more than half of participants using them exclusively and a smaller fraction combining OTC and prescription options. In contrast, the HeAD-US cohort demonstrated a higher adoption of migraine-specific therapies, such as gepants and anti-CGRP monoclonal antibodies, reflecting recent advancements in treatment accessibility and the advantages of digital recruitment strategies. Approximately 85% of HeAD-US participants reported using acute treatments, with prescription medications such as triptans and newer gepants being prominently used, especially among individuals with chronic migraine. Additionally, the study highlighted a notable uptake of medical devices, reflecting their increasing availability and acceptance in recent years. These findings underscore the evolving treatment landscape and highlight the potential of digital health tools to engage individuals with more complex or treatment-resistant migraine.

Our results indicate that the efficacy of acute migraine treatments varies significantly depending on the type of medication used. While there was no significant difference in overall treatment effectiveness between participants on polytherapy and those on monotherapy, the type of prescription monotherapy emerged as a critical determinant of outcomes. Gepants demonstrated the highest treatment efficacy, with over half of the participants reporting effective relief, outperforming triptans, barbiturates, and opioids. Notably, the superior effectiveness of gepants was consistent across both episodic migraine (EM) and chronic migraine (CM) subgroups. This finding is particularly striking given that gepants are often prescribed to individuals who have already failed first-line therapies, such as OTC medications or triptans, and who may therefore present with more complex or advanced disease. In contrast, barbiturates and opioids showed significantly lower efficacy rates, underscoring the need to prioritize evidence-based, migraine-specific treatments like gepants and triptans in clinical practice. These results emphasize the importance of tailoring acute migraine management strategies to individual patient needs while focusing on the most effective and well-tolerated options to improve outcomes.

The HeAD-US study has several limitations that should be acknowledged. First, the digital recruitment methodology may have introduced selection bias, as participants were likely more tech-savvy, health-conscious, and actively engaged in their migraine care, potentially overrepresenting individuals with more severe or treatment-resistant disease. Second, the reliance on self-reported data for medication use and treatment outcomes introduces the possibility of recall bias or misreporting, as there was no objective verification of the reported information. Third, the study may underrepresent certain demographics, such as older adults, individuals with limited digital access, and those from lower socioeconomic backgrounds. Therefore, our findings may not be generalizability to the broader migraine population. Lastly, as the current study is cross-sectional. It does not provide insights into long-term treatment efficacy, adherence, or changes in treatment patterns over time. To address this limitation, we are planning to expand the study into a longitudinal design, which will allow us to capture the dynamics of treatment responses and adherence over time.

The HeAD-US study provides valuable insights into the real-world diagnosis, treatment patterns, and outcomes for migraine patients using a digital health platform. With a large and diverse cohort, the study highlights significant advances in the availability and adoption of newer migraine-specific treatments, such as gepants and anti-CGRP monoclonal antibodies, while also underscoring persistent gaps in care, including the underuse of both acute and preventive therapies. The findings demonstrate the potential of digital platforms for large-scale, real-world data collection, which can help address critical knowledge gaps in migraine management and inform clinical practice. However, limitations such as selection bias, reliance on self-reported data, and the cross-sectional design indicate the need for cautious interpretation and further research. As the study expands to a longitudinal design, it promises to provide deeper insights into long-term treatment efficacy, adherence, and evolving patient needs, paving the way for improved, evidence-based approaches to migraine care.

## Data Availability

All data produced in the present study are available upon reasonable request to the authors

## References

1. Collaborators GA. Global, regional, and national burden of diseases and injuries for adults 70 years and older: systematic analysis for the Global Burden of Disease 2019 Study. bmj 2022;376.

2. Eigenbrodt AK, Ashina H, Khan S, et al. Diagnosis and management of migraine in ten steps. Nature Reviews Neurology 2021;17:501–514.

3. Awad A, Trenfield SJ, Pollard TD, et al. Connected healthcare: Improving patient care using digital health technologies. Advanced Drug Delivery Reviews 2021;178:113958.

4. Lipton RB, Munjal S, Alam A, et al. Migraine in America Symptoms and Treatment (MAST) study: baseline study methods, treatment patterns, and gender differences. Headache: The Journal of Head and Face Pain 2018;58:1408–1426.

5. Adams AM, Serrano D, Buse DC, et al. The impact of chronic migraine: The Chronic Migraine Epidemiology and Outcomes (CaMEO) Study methods and baseline results. Cephalalgia 2015;35:563–578.

6. Diamond S, Bigal ME, Silberstein S, Loder E, Reed M, Lipton RB. Patterns of diagnosis and acute and preventive treatment for migraine in the United States: results from the American Migraine Prevalence and Prevention study: CME. Headache: The Journal of Head and Face Pain 2007;47:355–363.

7. Lipton RB, Nicholson RA, Reed ML, et al. Diagnosis, consultation, treatment, and impact of migraine in the US: results of the OVERCOME (US) study. Headache: The Journal of Head and Face Pain 2022;62:122–140.

8. Shewale AR, Poh W, Reed ML, et al. Ubrogepant users’ real-world experience: Patients on ubrogepant, characteristics and outcomes (UNIVERSE) study. Headache: The Journal of Head and Face Pain 2024.

9. Lipton RB, Contreras-De Lama J, Serrano D, et al. Real-world use of ubrogepant as acute treatment for migraine with an anti-calcitonin gene-related peptide monoclonal antibody: results from courage. Neurology and Therapy 2024;13:69–83.

10. Manack Adams A, Hutchinson S, Engstrom E, et al. Real-world effectiveness, satisfaction, and optimization of ubrogepant for the acute treatment of migraine in combination with onabotulinumtoxinA: results from the COURAGE Study. The Journal of Headache and Pain 2023;24:102.

11. Goadsby PJ, Constantin L, Ebel-Bitoun C, et al. Multinational descriptive analysis of the real-world burden of headache using the Migraine Buddy application. European Journal of Neurology 2021;28:4184–4193.

12. Chiang C-C, Fang X, Horvath Z, et al. Simultaneous comparisons of 25 acute migraine medications based on 10 million users’ self-reported records from a smartphone application. Neurology 2023;101:e2560–e2570.

13. Lipton RB, Diamond S, Reed M, Diamond ML, Stewart WF. Migraine diagnosis and treatment: results from the American Migraine Study II. Headache: The Journal of Head and Face Pain 2001;41:638–645.

14. Kroenke K, Spitzer RL, Williams JB, Löwe B. An ultra-brief screening scale for anxiety and depression: the PHQ–4. Psychosomatics 2009;50:613–621.

15. Ezzati A, Jiang J, Katz MJ, Sliwinski MJ, Zimmerman ME, Lipton RB. Validation of the Perceived Stress Scale in a community sample of older adults. International journal of geriatric psychiatry 2014;29:645–652.

16. Lipton R, Kolodner K, Bigal M, et al. Validity and reliability of the migraine-treatment optimization questionnaire. Cephalalgia 2009;29:751–759.

17. Olesen J. Headache classification committee of the international headache society (IHS) the international classification of headache disorders. Cephalalgia 2018;38:1–211.

